# Association between stroke and psychosis across four nationally representative psychiatric epidemiological studies

**DOI:** 10.1101/2021.11.01.21265640

**Authors:** Vaughan Bell, William Tamayo-Agudelo, Grace Revill, David Okai, Norman Poole

**Affiliations:** Research Department of Clinical, Educational and Health Psychology, University College London, UK; South London and Maudsley NHS Foundation Trust, London, UK; Universidad Cooperativa de Colombia, Medellín, Colombia; Department of Neuropsychiatry, South London and Maudsley NHS Foundation Trust, London, UK; Department of Neuropsychiatry, South West London and St George’s Mental Health NHS Trust, London, UK

**Keywords:** Stroke, psychotic, delusions, hallucinations, neuropsychiatry

## Abstract

**Background:** Both stroke and psychosis are independently associated with high levels of disability. However, psychosis in the context of stroke has been under-researched. To date, there are no general population studies on their joint prevalence and association.

**Aims:** To estimate the joint prevalence of stroke and psychosis and their statistical association using nationally representative psychiatric epidemiology studies from two high-income countries – United Kingdom and the United States – and two middle-income countries – Chile and Colombia, and, subsequently, in a combined countries dataset.

**Methods:** Prevalences were calculated with 95% confidence intervals. Statistical association between stroke and psychosis, and stroke and psychotic symptoms, was tested using regression models. Overall estimates were calculated using an individual participant level meta-analysis on the combined countries dataset. The analysis is available online as a computational notebook.

**Results:** The overall prevalence of probable psychosis in stroke was 3.81% [95% CIs 2.34 - 5.82] and stroke in probable psychosis was at 3.15% [95% CIs 1.94 - 4.83]. The adjusted association between stroke and probable psychosis was OR = 3.32 [95% CIs 2.05 - 5.38]. On the individual symptom level, paranoia, hallucinated voices, and thought passivity delusion were associated with stroke in the unadjusted and adjusted analyses.

**Conclusions:** Rates of association between psychosis and stroke suggest there is likely a high clinical need group who are under-researched and may be poorly served by existing services.

## Introduction

Stroke and active psychosis are independently considered to cause some of the highest levels of disability among health conditions, meaning their combination is likely to result in severely debilitating outcomes for affected individuals. To add to the complexity, initial evidence suggests that the most common treatment for psychosis in stroke-affected individuals (antipsychotics) may increase mortality in this patient group (Su et al., 2021).

Despite the clear need for better evidence to inform care for affected patients, the first systematic review, and indeed the first review, dedicated to stroke and psychosis was only published in 2018 and focused specifically on poststroke psychosis (Stangeland et al., 2018). However, it is important to note that the association between stroke and psychosis extends beyond cases of poststroke psychosis and also includes those who have a preceding history of psychosis and are later affected by stroke. Indeed, people with a diagnosis of schizophrenia are at an increased risk of stroke and recurrent stroke (Fleetwood et al., 2021). The main treatment for psychosis, antipsychotic medication, raises the risk of stroke with evidence for the causal role of metabolic syndrome and cardiac arrhythmias (Nielsen et al., 2021). Studies on predictors of cardiovascular events more generally in patients with schizophrenia also highlight the role of shared risk factors that may raise the risk of both conditions independently (Osborn et al., 2015).

One difficulty in estimating the level of association between stroke and psychosis is that, as far as we are aware, all existing estimates have been drawn from clinical studies rather than population studies, meaning it is not clear to what extent estimates might be affected by selection biases, particularly referral bias. For example, all studies used to estimate prevalence of delusions and hallucinations in stroke patients included in the meta-analysis reported by Stangeland et al. (2018) were drawn from hospitalised stroke patients, potentially oversampling patients with the highest levels of disability. Indeed, hospitalisation referral biases have been evidenced for both stroke (Appelros et al., 2003) and psychosis (Sipos et al., 2001).

These limitations are particularly important when trying to estimate the association between stroke and psychosis in low- and middle-income countries where specialised stroke care may be less available. Indeed, low- and middle-income countries proportionally show the highest levels of stroke incidence and poor outcome (Feigin et al., 2009; Johnson et al., 2019), raising the possibility of whether stroke and psychosis might be a more frequent combination in developing countries.

Consequently, in this study, we identified epidemiological studies that recorded both stroke and psychosis from four countries and aimed to estimate the joint prevalence of stroke and psychosis and their statistical association. These included two high-income countries – the United Kingdom and the United States – and two middle-income countries – Chile and Colombia. Each of these countries have completed nationally representative psychiatric epidemiology studies that included structured assessments of psychotic disorders and / or psychotic symptoms, as well as measures of the participants’ health, including stroke status. We subsequently combined all national datasets into a single dataset to conduct an individual participant-level meta-analysis to estimate the overall prevalence and association between stroke and psychosis across all four countries.

## Methods

### Datasets

We used four nationally representative psychiatric epidemiological studies that recorded presence of both stroke and psychosis from the United Kingdom (UK), the United States of America (US), Chile and Colombia. Matched variables from across the four datasets were also merged to create a single combined countries dataset to conduct an internal individual participant-level meta-analysis. The original datasets are described below:

#### Adult Psychiatric Morbidity Survey 2007 (England, United Kingdom)

The Adult Psychiatric Morbidity Survey (APMS) 2007 was a household survey that used a multi-stage stratified probability sampling to recruit participants. Using the English national postcode database, private households were identified, and any resident individual aged 16 years or over was invited to participate. If more than one individual aged over 16 years was resident, one adult was randomly chosen to ensure the same chance of being selected for all eligible individuals. Psychotic symptoms were measured using the Psychosis Screening Questionnaire (PSQ) (Bebbington and Nayani, 1995). The PSQ is a 20-item interview that measures the presence of symptoms of hypomania, thought interference, persecution, perceptual abnormalities, strange experiences and hallucinosis. Full details of the survey, sampling methods and consent procedure are reported in McManus et al. (2009).

#### Collaborative Psychiatric Epidemiology Surveys 2001-2003 (United States)

The Collaborative Psychiatric Epidemiology Surveys (CPES) consisted of the three nationally representative surveys of mental health in United States: the National Comorbidity Survey Replication, the National Study of American Life and the National Latino and Asian American Study of Mental Health. A two-component sampling method was used to recruit participants. The first involved a multistage stratified area probability design to derive a nationally representative household sample and the second involved high-density supplemental sampling to oversample specific ethnic groups (Afro-Caribbean, Chinese, Filipino, Vietnamese and Puerto Rican). Psychotic symptoms were measured using the World Health Organization Composite International Diagnostic Interview (WHO-CIDI) 3.0 Psychosis Screen (Kessler and Üstün, 2004) that measures the lifetime presence of six symptoms visual hallucinations, auditory hallucinations, thought insertion, thought control, delusions of reference, and persecutory delusions. Full details of the survey, sampling methods and consent procedure are given in Heeringa et al. (2004).

### National Mental Health Survey 2015 (Colombia)

The National Mental Health Survey (NMHS) 2015 (Encuesta Nacional de Salud Mental) was a national survey of Colombia completed by the Ministry of Health and Social Protection (Ministerio de Salud y Protección Social). Participants were recruited using multistage stratified sampling that involved stratifying the population by region, municipality, and geographical area. Neighbourhood blocks in urban areas, and municipalities in rural areas were selected and all households were contacted for participation. Psychotic symptoms were measured using the WHO Self Reporting Questionnaire 24 (SRQ-24) (Harding et al., 1980) which was deployed as an interview, rather than a self-completion questionnaire. The SRQ-24 measures the presence of four psychotic symptoms: persecutory delusion, grandiosity, thought interference, and auditory hallucinations. Full details of the survey, sampling methods and consent procedure are reported in Gómez-Restrepo et al. (2016).

### National Health Survey 2016-2017 (Chile)

The National Health Survey (NHS) 2016-2017 (Encuesta Nacional de Salud) was a national survey by the Chilean Ministry of Health of non-institutionalised individuals aged 15 years and older in households in urban and rural areas across 15 regions of Chile. Participants were identified using stratified multistage probability sampling. Psychotic symptoms were measured with the WHO-CIDI 3.0 Psychosis Screen (Kessler and Üstün, 2004). Full details of the survey, sampling methods and consent procedure are given in Ministerio de Salud (2017).

#### Ethics

The authors assert that all procedures contributing to this work comply with the ethical standards of the relevant national and institutional committees on human experimentation and with the Helsinki Declaration of 1975, as revised in 2008. This study is a secondary data analysis of datasets that exist in the public domain and ethics approval for this human study was waived by University College London Research Ethics Committee. As can be seen from Table 1, different countries’ studies had different lower ages for their definition of adults (from 16-18) but all participants were consented as adults and provided written informed consent for participation in the original studies.

**Table 1.** Descriptive statistics for national and combined datasets.

|  | *United Kingdom*<br>*(N=7403)* | *United States*<br>*(N=6082)* | *Colombia*<br>*(N=10870)* | *Chile*<br>*(N=3403)* | *Combined*<br>*(N=27758)* |
| --- | --- | --- | --- | --- | --- |
| *Age* |  |  |  |  |  |
| Mean (SD) | 51.1 (18.6) | 41.5 (15.8) | 43.4 (16.8) | 49.4 (17.9) | 45.8 (17.7) |
| Median | 50.0 | 39.0 | 42.0 | 50.0 | 44.0 |
| [Min, Max] | [16.0, 95.0] | [18.0, 99.0] | [18.0, 96.0] | [17.0, 98.0] | [16.0, 99.0] |
| *Sex* |  |  |  |  |  |
| Male | 3197 (43.2%) | 2747 (45.2%) | 4384 (40.3%) | 1226 (36.0%) | 11554 (41.6%) |
| Female | 4206 (56.8%) | 3335 (54.8%) | 6486 (59.7%) | 2177 (64.0%) | 16204 (58.4%) |
| *Highest level of education* |  |  |  |  |  |
| No / primary education | 2278 (30.8%) | 540 (8.9%) | 4007 (36.9%) | 766 (22.5%) | 7591 (27.3%) |
| Mid-teen high school | 2103 (28.4%) | 835 (13.7%) | 5152 (47.4%) | 895 (26.3%) | 8985 (32.4%) |
| Late teen high school | 938 (12.7%) | 3604 (59.3%) | 895 (8.2%) | 1040 (30.6%) | 6477 (23.3%) |
| College / university | 1916 (25.9%) | 1103 (18.1%) | 713 (6.6%) | 668 (19.6%) | 4400 (15.9%) |
| Missing | 168 (2.3%) | 0 (0%) | 103 (0.9%) | 34 (1.0%) | 305 (1.1%) |
| *Stroke* |  |  |  |  |  |
| Yes | 180 (2.4%) | 171 (2.8%) | 79 (0.7%) | 95 (2.8%) | 525 (1.9%) |
| No | 7223 (97.6%) | 5724 (94.1%) | 10791 (99.3%) | 3287 (96.6%) | 27025 (97.4%) |
| Missing | 0 (0%) | 187 (3.1%) | 0 (0%) | 21 (0.6%) | 208 (0.7%) |
| *Paranoia* |  |  |  |  |  |
| Yes | 569 (7.7%) | 68 (1.1%) | 2340 (21.5%) | 59 (1.7%) | 3036 (10.9%) |
| No | 6834 (92.3%) | 4926 (81.0%) | 8530 (78.5%) | 3304 (97.1%) | 23594 (85.0%) |
| Missing | 0 (0%) | 1088 (17.9%) | 0 (0%) | 40 (1.2%) | 1128 (4.1%) |
| *Hallucinated voices* |  |  |  |  |  |
| Yes | 68 (0.9%) | 293 (4.8%) | 362 (3.3%) | 152 (4.5%) | 875 (3.2%) |
| No | 7335 (99.1%) | 4702 (77.3%) | 10508 (96.7%) | 3211 (94.4%) | 25756 (92.8%) |
| Missing | 0 (0%) | 1087 (17.9%) | 0 (0%) | 40 (1.2%) | 1127 (4.1%) |
| *Passivity delusion* |  |  |  |  |  |
| Yes | 77 (1.0%) | 33 (0.5%) | 534 (4.9%) | 18 (0.5%) | 662 (2.4%) |
| No | 7326 (99.0%) | 4962 (81.6%) | 10336 (95.1%) | 3345 (98.3%) | 25969 (93.6%) |
| Missing | 0 (0%) | 1087 (17.9%) | 0 (0%) | 40 (1.2%) | 1127 (4.1%) |
| *Probable Psychosis* |  |  |  |  |  |
| Yes | 66 (0.9%) | 36 (0.6%) | 505 (4.6%) | 27 (0.8%) | 634 (2.3%) |
| No | 7337 (99.1%) | 6046 (99.4%) | 10365 (95.4%) | 3376 (99.2%) | 27124 (97.7%) |

#### Variable coding and missing data management

Symptom data was recoded to represent strong evidence for the presence of symptoms. Where there was an ambiguous response data in response to interview questions about psychotic symptoms (‘Unsure’ responses in UK Adult Psychiatric Morbidity Survey; ‘don’t know’ or ‘didn’t respond’ responses in Chile National Health Survey) these were recoded as absent. Ambiguous responses were present at low rates: 0.4% of responses in the UK Adult Psychiatric Morbidity Survey data and 0.1% of the Chile National Health Survey data.

In the UK Adult Psychiatric Morbidity Survey, psychosis and stroke data was only collected after the participant responded ‘yes’ to initial screening questions, meaning missing data represented questions being intentionally not asked and therefore data was not missing at random. Consequently, the presence of stroke or psychotic symptoms was coded as not present for ‘no’ or ‘item not applicable’ data.

Missing data as originally present in the dataset is reported in Table 1. Notably, the majority of variables with missing data have low levels of missingness, below the threshold of 5% considered to be likely to bias estimates (Jakobsen et al., 2017). However, the psychotic symptom data from the US Collaborative Psychiatric Epidemiology Surveys data showed higher levels of missingness (approximately 18%). Random forest missing data imputation is highly reliable in reducing bias in estimates (Shah et al., 2014). Consequently, the missing psychotic symptoms values were imputed using the *missForest* package for *R*. No additional instances of symptoms were imputed for the Collaborative Psychiatric Epidemiology Surveys data and so all missing variables were coded as ‘not present’.

### Variables

#### Exposure

In all studies, individuals were asked to report if they had been diagnosed with a stroke by a doctor during the structured health assessment. This was used as the primary exposure variable in regression analyses.

#### Outcome

Due to the use of differing psychosis measures across studies, we extracted symptom-level items from interviews that measured the presence of the following symptoms across all four studies: i) paranoia, ii) hallucinated voices, and iii) thought passivity delusion.

There was no consistent metric for probable psychosis across surveys. The Adult Psychiatric Morbidity Survey and the Collaborative Psychiatric Epidemiology Surveys had inconsistent criteria (the former was based solely on symptom screening, the latter included service use – i.e. antipsychotic, hospital admission) and the other surveys did not code for this category. Therefore, we created a standardised criteria for the category of ‘probable psychosis’ that was coded when any two psychotic symptoms were present. Consequently, probable psychosis was coded when a participant reported at least one delusion-like belief along with hallucinated voices, or at least two delusion-like beliefs at least one of which was a thought-passivity experience.

#### Potential confounders

A graph analysis that mapped major evidenced risk factors between stroke and psychosis (Rohrer, 2018) indicated that the total effect between stroke and psychosis could not be estimated by covariate adjustment, largely due to the reciprocal causal relationship between stroke and psychosis and the role of alcohol and smoking, which act as mediators.

However, we selected a minimal group of potential confounders that were most likely to represent pre-onset risk factors for both stroke and psychosis, namely age, sex, and highest level of education, to help refine the estimate. Age and sex are independent predictors of stroke (Avan et al., 2019) and psychosis (Jongsma et al., 2019) before onset. There was no consistent measure of pre-onset socioeconomic status in all four studies. However, highest level of education, which correlates strongly with socioeconomic status and is frequently used as a component measure of it (Cox et al., 2006), is a pre-onset predictor of both stroke (Addo et al., 2012) and psychosis (Hakulinen et al., 2019) was included. Highest educational attainment was recoded across studies to a consistent coding of ‘no or primary education only’, ‘mid-teen high school’, ‘late teen high school’, and ‘college / university’.

Both alcohol use and smoking are likely to be independent risk factors for both stroke (Pan et al., 2019; Reynolds et al., 2003) and psychosis (Jørgensen et al., 2018; Mustonen et al., 2018). However, there is also strong evidence for psychosis as a causal risk factor for smoking and alcohol use (Hartz et al., 2014) potentially indicating its additional role as a mediating factor. Furthermore, smoking and alcohol intake were only measured contemporaneously in the studies that reported them. Given these issues, alcohol and smoking were not included as potential confounders in the analysis.

### Analysis

All analysis was conducted using *R* version 4.0.3 and the full code and output for the analysis is available in the format of a Jupyter Notebook, a document that combines both code and the output in a form that can be re-run and reproduced. All analysis code is available at the following link: https://github.com/vaughanbell/stroke-psychosis-national-epi-analysis

#### Prevalence

We calculated the prevalence of stroke, prevalence of probable psychosis, prevalence of probable psychosis in people with stroke, and prevalence of stroke in people with probable psychosis using the *epiR* package. Prevalence and 95% confidence intervals were calculated for each of these for each individual national study and meta-analytically, using the combined countries dataset.

#### Association between stroke and psychosis

At the national dataset level, we used logistic regression models to estimate the association between stroke, probable psychosis, and individual psychotic symptoms (paranoia, hallucinated voices, passivity delusion). We first estimated the unadjusted association and then the adjusted association – adjusted for sex, age, and highest level of education. Survey weights were of an incompatible format between datasets and so were not included in the analysis.

#### Individual participant-level meta-analysis

We completed an internal meta-analysis of individual participant data by additionally conducting the prevalence and regression analyses on the combined countries dataset. Following recommendations from Riley et al.(Riley et al., 2010), when completing the regression analyses we accounted for potential clustering of participants within studies by using multi-level regression models where country was added as a random effect.

## Results

Descriptive statistics for each national survey and the combined dataset are reported in Table 1. The demographic profile was broadly similar across national surveys.

### Prevalence

Table 2 displays the calculated prevalence with 95% confidence intervals for stroke, probable psychosis, probable psychosis in stroke, and stroke in probable psychosis. The larger estimates of prevalence within national surveys tend to be accompanied by wider confidence intervals although the estimates for the combined countries dataset have consistently narrower confidence intervals, suggesting more reliable estimates.

**Table 2.** Prevalence and 95% confidence intervals (CIs) for stroke, probable psychosis, probable psychosis in stroke, and stroke in probable psychosis across the four nations and combined country datasets

|  | *Prevalence (95% CIs)* |  |  |  |
| --- | --- | --- | --- | --- |
|  | *Stroke in Total Population* | *Probable Psychosis in Total Population* | *Probable Psychosis in Stroke Population* | *Stroke in Probable Psychosis Population* |
| United Kingdom | 2.43% (2.09 - 2.81) | 0.89% (0.69 - 1.13) | 1.11% (0.13 - 3.96) | 3.03% (0.37 - 10.52) |
| United States | 2.90% (2.49 - 3.36) | 0.59% (0.41 - 0.82) | 3.51% (1.30 - 7.48) | 16.67% (6.37 - 32.81) |
| Colombia | 0.73% (0.58 - 0.90) | 4.65% (4.26 - 5.06) | 13.92% (7.16 - 23.55) | 2.18% (1.09 - 3.86) |
| Chile | 2.81% (2.28 - 3.42) | 0.79% (0.52 - 1.15) | 1.05% (0.03 - 5.73) | 3.70% (0.09 - 18.97) |
| Combined meta-analytic estimate | 1.91% (1.75 - 2.07) | 2.28% (2.11 - 2.47) | 3.81% (2.34 - 5.82) | 3.15% (1.94 - 4.83) |

### Association and adjusted associations between stroke and psychosis

Unadjusted associations between stroke and psychosis alongside associations adjusted for potential confounders are reported in Table 3. In addition, Table 4 reports unadjusted and adjusted associations for specific symptoms of psychosis.

**Table 3.** Results of unadjusted and adjusted logistic regression analysis reporting associations between stroke and probable psychosis with 95% confidence intervals (CIs)

|  | *Odds Ratios (95% CIs)* |  |
| --- | --- | --- |
|  | *Unadjusted* | *Adjusted* |
| *Probable Psychosis* |  |  |
| United Kingdom | 1.26 (0.31 - 5.17) | 1.11 (0.15 - 8.26) |
| United States | 6.90 (2.83 - 16.81) | 6.22 (2.52 - 15.35) |
| Colombia | 3.37 (1.77 - 6.42) | 4.04 (2.10 - 7.78) |
| Chile | 1.33 (0.18 - 9.94) | 1.33 (0.18 - 10.09) |
| Combined meta-analytic estimate | 3.06 (1.93 - 4.85) | 3.32 (2.05 - 5.38) |

**Table 4.** Results of unadjusted and adjusted logistic regression analysis reporting associations between stroke and psychotic symptoms with 95% confidence intervals (CIs)

|  | *Odds Ratios (95% CIs)* |  |
| --- | --- | --- |
|  | Unadjusted | Adjusted |
| *Paranoia* |  |  |
| United Kingdom | 0.63 (0.32 - 1.23) | 0.99 (0.46 - 2.16) |
| United States | 2.71 (1.07 - 6.82) | 2.50 (0.99 - 6.34) |
| Colombia | 2.01 (1.27 - 3.20) | 2.34 (1.46 - 3.74) |
| Chile | 1.92 (0.59 - 6.26) | 1.71 (0.52 - 5.66) |
| Combined meta-analytic estimate | 1.38 (1.00 - 1.90) | 1.66 (1.19 - 2.32) |
| *Hallucinated voices* |  |  |
| United Kingdom | 0.60 (0.08 - 4.32) | 0.85 (0.11 - 6.29) |
| United States | 2.33 (1.41 - 3.86) | 2.03 (1.22 - 3.38) |
| Colombia | 4.30 (2.20 - 8.41) | 4.06 (2.05 - 8.03) |
| Chile | 0.71 (0.22 - 2.26) | 0.71 (0.22 - 2.29) |
| Combined meta-analytic estimate | 2.01 (1.39 - 2.90) | 1.89 (1.30 - 2.74) |
| *Thought passivity delusion* |  |  |
| United Kingdom | 1.07 (0.26 - 4.40) | 0.67 (0.09 - 4.94) |
| United States | 2.17 (0.52 - 9.16) | 2.12 (0.50 - 9.00) |
| Colombia | 3.52 (1.89 - 6.55) | 4.27 (2.27 - 8.03) |
| Chile | 2.09 (0.28 - 15.87) | 2.17 (0.28 - 16.95) |
| Combined meta-analytic estimate | 2.51 (1.52 - 4.14) | 2.68 (1.59 - 4.52) |

In the regression analyses, stroke does not reliably predict probable psychosis in the United Kingdom and Chile, although it is a reliable predictor in the United States, Colombia, and in the combined countries dataset.

There is variation in the extent to which stroke is reliably associated with individual psychotic symptom measures across countries, although stroke is reliably associated with all symptoms in the combined countries dataset.

## Discussion

We report the joint prevalences of stroke and probable psychosis across four nationally representative epidemiological studies, and subsequently the association between stroke and probable psychosis after adjustment for potential confounders. We subsequently calculated overall estimates from a combined countries dataset using individual participant level meta-analysis. We found that the prevalence of probable psychosis in people with stroke ranged from 1.05% (Chile) to 13.92% (Colombia) with the prevalence from the combined countries dataset estimated at 3.81%. Conversely, the prevalence of stroke in people with probable psychosis ranged from 2.18% (Colombia) to 16.67% (US) with the combined countries prevalence estimated at 3.15%. However, the larger estimates were accompanied by wide confidence intervals and are less likely to be accurate estimates of true population prevalence. Estimates for the adjusted association between stroke and probable psychosis ranged from an odds ratio of 1.11 (95% CI 0.15 - 8.26) in the UK to an odds ratio of 6.22 in the US (95% CIs 2.52 - 15.35) with the association from the combined dataset estimated at 3.32 (95% CIs 2.05 - 5.38). We also examined the association between stroke and paranoia, hallucinated voices, and thought passivity delusion, and although we found significant variation in the reliability and strength of association across countries, all three psychotic symptoms were associated with stroke in the unadjusted and adjusted analyses in the combined countries dataset.

This evidence suggests a relatively high co-prevalence of stroke and psychosis with approximately 1 in 26 people with stroke having probable psychosis and 1 in 32 of people with probable psychosis having stroke across the combined countries dataset. This is despite the fact that psychosis has often been described as a “rare” complication of stroke in the literature and has mostly been reported as single case studies or case series. The estimate here is broadly in line with previous estimates of single psychotic symptoms in patients with stroke with meta-analytic estimates (admittedly from a small number of studies) suggesting a delusion prevalence of 4.67% and hallucination prevalence of 5.05% (Stangeland et al., 2018).

We also note that the majority of research in this area has focused on poststroke psychosis that likely contributes only a proportion of the co-prevalence of stroke and psychosis. As psychosis, and treatment for psychosis, is a risk factor for later stroke (Li et al., 2014; Marto et al., 2021), stroke in patients with a preceding history of psychosis is also likely to be an important contributory factor to co-prevalence. We also note here that initial studies report that patients with psychosis who later experience stroke have worse outcomes and are less likely to receive equitable care (Kisely et al., 2009) including timely invasive interventions (Nielsen et al., 2021). Taken together, this evidence suggests that stroke and psychosis may be highly disabling, but is under-recognised and likely under-served by existing services.

It is important to note that significant international variability was found in the estimates of association – either through calculating prevalence or odds ratios – between stroke and psychosis. Given the variability of measures used to measure psychotic symptoms within countries, one key question is the extent to which these estimates are being affected by characteristics of the measures, versus the extent to which the prevalence of psychotic symptoms and their association with stroke varies between countries.

We note two standout prevalence figures. A prevalence of probable psychosis in stroke of 13.92% in Colombia and a prevalence of stroke in probable psychosis of 16.67% in the US. Both of these figures have wide confidence intervals and the accompanying alternative prevalences (stroke in probable psychosis in Colombia, and probable psychosis in stroke in the US) are within the more typical ranges internationally. This suggests they may be less accurate estimates of the true prevalence. However, it remains challenging to separate measurement error from population-specific risk factors that contribute to these larger figures given the cross-sectional nature of the data.

For example, the estimated rate of probable psychosis in the total population is markedly higher in Colombia, which also has the highest estimated rate of probable psychosis in stroke. We note here that several factors may be important in influencing this outcome. The Colombia National Mental Health Survey used the WHO Self Reporting Questionnaire 24 (Harding et al., 1980) which although was deployed in an interview format, solely relied on participant answers without any judgment from the trained interviewers regarding the likelihood of the answer representing a symptom. Although self-report questionnaires for psychotic symptoms show broad agreement with interview measures they may over-report milder symptoms. We also note that, of the four countries included in this analysis, Colombia has the highest rate, and an internationally high rate, of violence and victimisation as well as experience of a long-running armed conflict. This likely contributes both to the over-rating of the paranoia item in terms of it measuring genuine threat rather than the exaggerated perception of threat, as well as likely increasing the rate of genuine paranoia as psychopathology, due to the fact that violent victimisation is associated with a higher risk of subsequent psychosis.

The high prevalence, wide-confidence-interval estimate of stroke in probable psychosis prevalence in the US, likely reflects the fact that it has the highest prevalence of stroke among countries along with the lowest prevalence of psychosis reported here. There is some evidence that stroke prevalence may be slightly over-estimated in this study: 2.9% US stroke prevalence reported here vs 2.6% reported by the Centers for Disease Control and Prevention (CDC) for non-institutionalised US adults in 2005 (Centers for Disease Control and Prevention, 2007). It is possible psychosis prevalence was slightly under-estimated. The Global Burden of Disease study reported higher rates of schizophrenia in the US compared to the three other countries reported here (Charlson et al., 2018), despite it having the lowest rate of psychosis estimated in this study.

One potential way to interpret this data is to compare the extent to which the prevalence of probable psychosis used in this study compares to the prevalence of psychosis phenotypes. Here, our probable psychosis category as applied to the UK, US and Chile surveys are more likely to be measuring a narrow psychosis phenotype more akin to psychotic disorder, whereas in Colombia, it is more likely to be measuring a broader psychosis phenotype of psychotic experiences (van Os et al., 2009). However, it is also worth noting that in a recent systematic review of poststroke psychosis, delusional disorder, typically involving a single isolated delusion, was the most commonly reported psychosis in post-stroke cases – albeit from a relatively poor quality evidence base (Stangeland et al., 2018). The probable psychosis criteria used in this study would have excluded these cases, indicating that this may have underestimated the full prevalence of psychosis.

An additional factor is the extent to which stroke, psychosis, and their possible combination may be under-reported in community epidemiological studies due to a case ascertainment bias – in that those with more severe difficulties are less likely or less able to participate. Aked et al. (2020) compared stroke ascertainment between a community epidemiological study and a clinical register and reported that the community study was more likely to detect milder strokes but was equally likely to detect more severe cases. Nevertheless, the data used in the present study was from psychiatric epidemiology studies that require active participation in an extensive interview. Given this, it is likely that this may have led to an under-representation of more severe stroke or communication-impairing strokes in the dataset, and potentially, cases with more severe disability caused by a combination of stroke and psychosis.

We also note here that stroke was measured in all surveys by an interview item asking whether the person had been diagnosed with stroke by a doctor. Self-reported stroke has been found to have a consistently high negative predictive value but a variable positive predictive value (22–87%) with the misreporting of transient ischaemic attacks for stroke likely to be a major contributor to false positive reporting (Woodfield et al., 2015). However, the measure used in this study was not self-reported stroke per se, but self-report of doctor-diagnosed stroke. As far as we are aware, the only study we know that has examined the accuracy of this specific method of reporting stroke was Walker et al. (1998) where self-report of doctor-diagnosed stroke had a positive predictive value of 0.89 with the majority of false positive being reports of transient ischaemic attacks. Hence, the measure included in this study is most likely to represent both stroke proper and transient ischaemic attack. In addition, this measure is likely to be affected by the number of doctors available to diagnose stroke. This may underestimate prevalence in lower income countries where healthcare may be less accessible or inaccessible, or more likely to be carried out by non-physician healthcare professionals, particularly in remote or rural areas. The lack of detail beyond the presence of absence of stroke also means it is not possible to make inferences regarding the relationship between stroke type, severity, location, recurrence and psychosis. Accordingly, studies using formal diagnosis and additional data on stroke characteristics are needed to ensure the highest accuracy of estimates and associations.

We also note that the differing availability of mental health services could affect the prevalence of psychotic symptoms because although each study measured psychotic symptoms by interviewing the participant directly, effective available treatment could reduce the presence of symptoms.

There are additional limitations that should be noted. Some potentially useful covariates could not be included because they were not measured in all datasets. One is the extent to which the findings provide a guide to future stroke and psychosis prevalence given the improving stroke survival rates in high-income countries, largely due to improvement in acute stroke care (Joundi et al., 2021). Given the high rates of stroke risk factors in individuals with pre-existing psychosis, we suggest this will increase the rates of post-psychosis stroke due to better survival rates, and it is possible that this may increase the rates of post-stroke psychosis, although the relationship between stroke severity and psychosis risk is still poorly understood. We also note the increasing incidence of stroke in the young globally (Boot et al., 2020), potentially changing the risk profile of stroke, and comorbid stroke and psychosis.

Due to the fact that this study uses cross-sectional data, the extent to which the association between stroke and psychosis consisted of poststroke psychosis, versus people with psychosis who later experienced stroke, was impossible to determine. Longitudinal studies will be needed to address these key questions, and we note here that longitudinal studies examining to what extent psychosis occurs post-stroke and to what extent stroke occurs post-psychosis, but crucially, measured within the same cohort, are likely to be important in addressing these key issues. This information is clearly important in developing both preventative healthcare and understanding how specific services (specifically psychiatry and neurology) should prioritise treatment and referral, given that the order of which psychosis or stroke occurs is likely to determine which service a patient has first contact with.

We also suggest that involvement of more integrated psychological medicine services in stroke services including both psychiatry and psychology is likely to be important, as is prioritising management of stroke risk factors in patients with psychosis (Cooper et al., 2016). In addition, psychiatrists should be aware of the signs and symptoms of stroke, including apparently ‘silent stroke’, and be aware of timely referral pathways to specialist stroke services in their area.

In conclusion, we report the first study on the association of stroke and psychosis in the general population that examines the co-prevalence and association within four countries: the US, UK, Colombia and Chile. We note that the conditions co-occur more frequently than has previously been assumed and there remains a marked lack of research in this area. This is a particular priority given the potentially high need of these patient groups and the potential avoidance of stroke if risk factors are appropriately managed. Future research needs to involve standardised diagnostic assessments and longitudinal studies to determine the extent to which stroke and psychosis appear in specific causal sequences.

## Data Availability

This study is a secondary analysis of existing datasets and so produced no new raw data.

https://github.com/vaughanbell/stroke-psychosis-national-epi-analysis

## Acknowledgements

None

## Notes

**Funding** Funding Statement: This work was supported by a Medical Research Council Doctoral Training Partnership awarded to GR.

### Competing Interest Statement

The authors have declared no competing interest.

### Funding Statement

This study did not receive any funding

### Author Declarations

This study is a secondary analysis of existing datasets made public by the institutions that collected the data. The relevant ethical approvals are: Adult Psychiatric Morbidity Survey 2007 (UK) The Research Ethics Committee of the Royal Free Hospital and Medical School gave ethical approval for the original study. Collaborative Psychiatric Epidemiology Surveys 2001-2003 (US) The Institutional Review Board of the University of Michigan gave ethical approval for the original study. National Mental Health Survey 2015 (Colombia) The review boards of the the Pontificia Universidad Javeriana and Colciencias gave ethical approval for this study. National Health Survey 2016-2017 (Chile) The ethics committee of the Escuela de Medicina de la Pontificia Universidad Catolica de Chile have ethical approval for this study. Links to all datasets are listed on the github page of the present study: https://github.com/vaughanbell/stroke-psychosis-national-epi-analysis

### Summary of Updates

Include better description of individual participant level meta-analysis

